# A cross-sectional analysis of factors associated with the development of refeeding syndrome in children 0 – 59 months diagnosed with severe acute malnutrition in a South African setting

**DOI:** 10.1101/2022.06.03.22275953

**Authors:** Natalie Heydenrych, Tim De Maayer, Mariette Nel, Louise van den Berg

**Author notes:** **Correspondence to:** Louise van den Berg. These authors contributed equally to this work.

## Abstract

**Background:** Refeeding syndrome (RFS) is a life-threatening, underdiagnosed, and under-researched complication in treating children with severe acute malnutrition (SAM). This study aimed to determine the incidence and onset of RFS and identify biochemical abnormalities, clinical signs, and complications associated with RFS development in children 0–59 months treated in a South African public hospital setting.

**Methods:** A retrospective cohort study was performed on hospital files of children diagnosed with SAM at Rahima Moosa Mother and Child Hospital, Johannesburg, from 1/10/2014 to 31/12/2018. A total of 148 files could be retrieved from the hospital archives. The diagnosis of SAM based on the World Health Organization definition was confirmed in 126 of these children, and they were included in the study. The onset of RFS among the children included in the study was diagnosed based on published criteria for RFS. Children who developed RFS and those who did not were compared concerning biochemistry and clinical signs and symptoms on admission.

**Results:** The median age of the 126 children (63% male) with confirmed SAM was 34 months (IQR: 26.0 to 48.4 months). The mortality rate was 18.2%. Of these children, 8.7% were retrospectively diagnosed as having developed RFS during their recorded hospital stay, despite implementing the WHO treatment guidelines for SAM. A significantly higher percentage of the children that developed RFS presented on admission with hypophosphatemia (p=0.04), severe hypokalemia (p=0.0005), hyponatremia (p=0.004), an international normalized ratio (INR) of above 1.7 (p=0.049), diarrhea (p=0.04), dehydration (p=0.02) and urinary tract infection (UTI) (p=0.04) than those that did not. Edema was more prevalent on admission in children who developed RFS than those who did not (63.6% vs 39.1%), though the difference was not statistically significant (p=0.20). Children who developed RFS stayed in hospital significantly longer than those who did not (18 vs 12 days) (p=0.003).

**Conclusion:** In this population of children with SAM treated in a South African public hospital setting, the presence on hospital admission of low levels of electrolytes, elevated INR, dehydration, diarrhea, and UTI was significantly associated with developing RFS. Recognizing these as possible red flags for developing RFS in children admitted with SAM might contribute to improved outcomes and needs further investigation.

## Introduction

Severe acute malnutrition is a term used by the World Health Organization in children under five years of age to indicate severe undernutrition and is diagnosed by the occurrence of either severe wasting, i.e., weight-for-length/height <-3 standard deviation (SD) on the WHO growth standards; a mid-upper arm circumference (MUAC) <11.5 cm in children between 6 and 59 months; or the presence of bilateral pitting edema [1]. According to the WHO, following the WHO management guidelines for SAM may reduce the case fatality rate to below 10% [2]. However, Tickell & Denno highlighted that these guidelines were developed mainly based on expert opinion rather than scientific evidence and that additional research is vital to improving the guidelines and children’s outcomes [2].

Since World War II, it has been known that initiating feeds in a starved patient may potentially contribute to their fatality [3,4]. When feeding is initiated in an individual after starvation, a sudden shift from catabolism to anabolism and a switch back to carbohydrate metabolism causes blood glucose and insulin levels to rise and phosphate, potassium, and magnesium to rise and shift into the intracellular space [5]. Fluid retention may also increase the extracellular fluid volume [5]. Thus, the syndrome is characterized by a drop in blood phosphate to below normal levels (hypophosphatemia) within five days [6] of initiating nutritional therapy. Clinical symptoms range from mild to severe and may be associated with cardiac arrhythmias, cardiac failure, renal failure, and death [6]. Preventing the onset of RFS in patients treated with SAM may improve patient outcomes and reduce mortality. However, most RFS studies focus on adults, children with anorexia, and those in intensive settings. Very little research has focused on RFS in the context of SAM [7–11], while guidelines for treating SAM [1] and RFS [5,7–9] have been developed independently. Although the WHO guidelines for the treatment of SAM include progressing feeding slowly [1], a lack of research in this field makes it difficult to determine whether this approach is optimal for preventing RFS in this vulnerable population.

In the African context, only two published studies thus far have reported RFS incidence in children admitted with SAM [4,7]. Mbethe & Mda reported on 104 children with SAM treated according to the WHO management protocol in a teaching hospital in Gauteng Province [4], while Okinyi studied 160 children with SAM in a Kenyan public hospital after initiating feeding with F-75 [7]. Considering the paucity of data, this study aimed to determine the incidence of RFS and explore associated admission data that might indicate risk factors for the development of RFS in South African children aged 0 – 59 months diagnosed with SAM and treated according to the WHO treatment guidelines in a public health setting.

## Methods

Ethics approval was obtained from the Health Sciences Research Ethics Committee of the University of the Free State (UFS-HSD2018/0154/2602), and permission to do the study at the Rahima Moosa Mother and Child Hospital was granted by the Gauteng Department of Health and the hospital management.

### Study design, setting, and study population

A retrospective cohort study of retrievable hospital files of children aged 0 – 59 months admitted with SAM to Rahima Moosa Mother and Child Hospital, Coronationville, Johannesburg, South Africa, from 1 October 2014 to 31 December 2018, was conducted.

According to electronic statistics kept by the Dietetics Department of the hospital, 592 children with SAM were admitted during the period under review; however, only 148 of the hospital files of these children (25% of the identified total) could be retrieved from the Hospital Archives Department. The diagnosis of SAM was verified based on the World Health Organization definition [1] according to the weight, length, MUAC, and presence of edema captured in these files. The diagnosis of SAM could be confirmed for 126 children who were then eligible for inclusion in the current study. Thus, the sample represented 21% of the children admitted to the hospital with SAM during the time under review.

### Data collection

Anthropometry (growth indicators), age, gender, ethnicity, country of origin, and clinical outcomes were recorded from the hospital files. Biochemical values associated with SAM, RFS, or prognosis were recorded; these included blood levels of electrolytes and minerals (phosphorus, potassium, magnesium, calcium, and sodium), indicators of kidney function (urea and creatinine), inflammation and metabolic stress (C-reactive protein (CRP) and albumin), hemoglobin, coagulation (platelets and international normalized ratio (INR), and indicators of liver function (total bilirubin, alanine aminotransferase (ALT), alkaline phosphatase (ALP), gamma-glutamyl transferase (GGT) and aspartate aminotransaminase (AST). Values were interpreted according to published cut-offs. All clinical signs and medical complications recorded in the admission files were documented.

### Data analysis

The onset of RFS among the children included in the study was retrospectively diagnosed if a drop in blood phosphate levels by ≥0.16 mmol/L to below 0.65 mmol/L occurred after feeding was initiated. There is no universally accepted definition of RFS [6], but according to a recent systematic review[3], this definition by Marik and Bedigian [17] is one of the most frequently used. The sample was stratified into children who developed RFS (the RFS-positive group) and those who did not (the RFS-negative group). Recorded variables were compared between the two groups.

### Statistical analysis

The data from the hospital files were captured in Microsoft Excel (2013) and analyzed using SAS® version 9.4, copyright© 2014 (SAS Institute Inc. SAS and all other SAS Institute Inc. product or service names are registered trademarks or trademarks of SAS Institute Inc., Cary, NC, USA.). Medians and percentiles were used to describe numerical data, and frequencies and percentages to describe categorical data. The groups were compared using contingency tables applying the Kruskal-Wallis test for numerical data and Chi-square or Fisher’s exact tests, as applicable, for categorical data. A value of p<0.05 was considered statistically significant.

## Results

The study included 126 children in whom the diagnosis of SAM could be retrospectively confirmed, of which 11 (8.7%) developed RFS after feeding was initiated. The children had a median age of 34.2 months (IQR: 26.0 to 48.4 months). The sample was stratified according to children that developed RFS (RFS-positive group) and those that did not (RFS-negative group). Most children were male (62.7%), and most were of African descent (95.2%) and from South Africa (64.3%) and Zimbabwe (21.4%) (Table 1). The demographical data did not differ significantly between the two groups (Table 1).

**Table 1:** Demographic data of the children, stratified according to the development of refeeding syndrome (RFS) (n=126)

| Variables and categories |  | All (n=126) |  | RFS-positive group (n=11) |  | RFS-negative group (n=115) |  | p-value<sup>1</sup> |
| --- | --- | --- | --- | --- | --- | --- | --- | --- |
|  |  | n | % | n | % | n | % |  |
| Gender (n=126) | Male | 79 | 62.7% | 7 | 63.6% | 72 | 62.6% | 1.00 |
|  | Female | 47 | 37.3% | 4 | 36.4% | 43 | 37.4% |  |
| Ethnicity (n=126) | White | 0 | - | 0 | - | 0 | - | 0.70 |
|  | African | 120 | 95.2% | 11 | 100% | 109 | 94.8% |  |
|  | Mixed race | 4 | 3.2% | 0 | - | 4 | 3.5% |  |
|  | Indian | 2 | 1.6% | 0 | - | 2 | 1.7% |  |
| Nationality (n=111) | South Africa | 72 | 64.3% | 5 | 55.6% | 67 | 65.1% | 1.00 |
|  | Zimbabwe | 24 | 21.4% | 3 | 33.3% | 21 | 20.4% |  |
|  | Mozambique | 8 | 7.1% | 1 | 11.1% | 7 | 6.8% |  |
|  | Malawi | 4 | 3.6% | 0 | - | 4 | 3.9% |  |
|  | Pakistan | 1 | 0.9% | 0 | - | 1 | 1.0% |  |
|  | India | 1 | 0.9% | 0 | - | 1 | 1.0% |  |
|  | Congo, DRC | 1 | 0.9% | 0 | - | 1 | 1.0% |  |

<sup>1</sup>p-value for the difference between the RSF-positive and RSF-negative groups; p<0.05 was considered statistically significant

The anthropometry, length of hospital stay, and mortality are summarized in Table 2. The RFS-positive group was younger, shorter, and weighed less than the RFS-negative group, but the differences were not statistically significant. MUAC was recorded for fewer than half of the children (n=60; 47.6%) and did not differ between RFS positive and negative groups. Edema was present in 63.6% and 39.1% (p=0.11), and severe wasting in 45.5% and 60.9% (p=0.32) of the RFS positive and RSF negative groups, respectively. The median duration of hospitalization was 12 days (IQR: 8 to 17 days). The median stay in hospital was significantly longer for the RFS-positive group than for the RFS-negative group (18 vs 12 days; p=0.003).

**Table 2:** Anthropometry and duration of hospital stay of the children, stratified according to the development of refeeding syndrome (RFS)

|  | All<br>(n=129) |  |  | RFS-positive group<br>(n=11) |  |  | RFS-negative group<br>(n=129) |  |  | p-value<sup>1</sup> |
| --- | --- | --- | --- | --- | --- | --- | --- | --- | --- | --- |
|  | n | Median | P25;P75 | n | Median | P25;P75 | n | Median | P25;P75 |  |
| Age on admission (months) | 126 | 34.2 | 26.0; 48.4 | 11 | 9.4 | 8.1; 17.8 | 115 | 12.8 | 7.7; 19.7 | 0.95 |
| Weight on admission (kg) | 126 | 6.3 | 5.3; 7.3 | 11 | 5.6 | 4.7; 7.3 | 115 | 6.4 | 5.3; 7.3 | 0.45 |
| Length/height on admission (cm) | 121 | 69.0 | 65.0; 77.0 | 10 | 67.5 | 62.0; 70.0 | 111 | 69.0 | 65.0; 77.5 | 0.46 |
| MUAC on admission (cm) | 60 | 11.1 | 10.4; 12.5 | 4 | 11.2 | 10.3; 12.7 | 56 | 11.1 | 10.4; 12.5 | 0.95 |
| Duration of hospitalization (days) | 125 | 12 | 8; 17 | 11 | 18 | 15; 27 | 114 | 12* | 7; 16 | 0.003* |
| Weight on discharge (kg) | 105 | 7.1 | 6.1; 8.3 | 9 | 6.9 | 6.0; 7.6 | 96 | 7.1 | 6.4; 8.3 | 0.75 |
| Weight gain during hospitalization (g/kg/day) | 105 | 0.6 | 0.2; 0.9 | 9 | 0.9 | 0.8; 1.0 | 96 | 0.5 | 0.2; 0.9 | 0.25 |
| Time until death (days) | 19 | 9 | 2; 15 | 2 | 14 | 9; 19 | 17 | 7 | 2; 15 | 0.29 |
| Inpatient mortality | 19 | 15.1 |  | 2 | 18.2 |  | 17 | 14.8 |  | 0.28 |

<sup>1</sup> p-value for the difference between the RFS-positive and RFS-negative groups; p<0.05 was considered statistically significant (\*)

Overall, children gained a median of 0.6 g/kg/day (IQR: 0.2 to 0.9 g/kg/day) during their hospital stay. Nineteen (19) children (15.1%) died in hospital, and the median time until death was nine days (IQR: 2 to 13 days). However, the mortality rate did not differ significantly between the RSF positive (n=2; 18.2%) and the RSF negative groups (n=17, 14.8%).

Table 3 summarizes the abnormal biochemical findings, showing that on admission, a significantly larger percentage of children in the RFS-positive group presented with hypophosphatemia (p=0.04) and severe hypokalemia (0.0005), hyponatremia (p=0.004), and elevated INR (p=0.049) than in the RFS negative group.

**Table 3:**
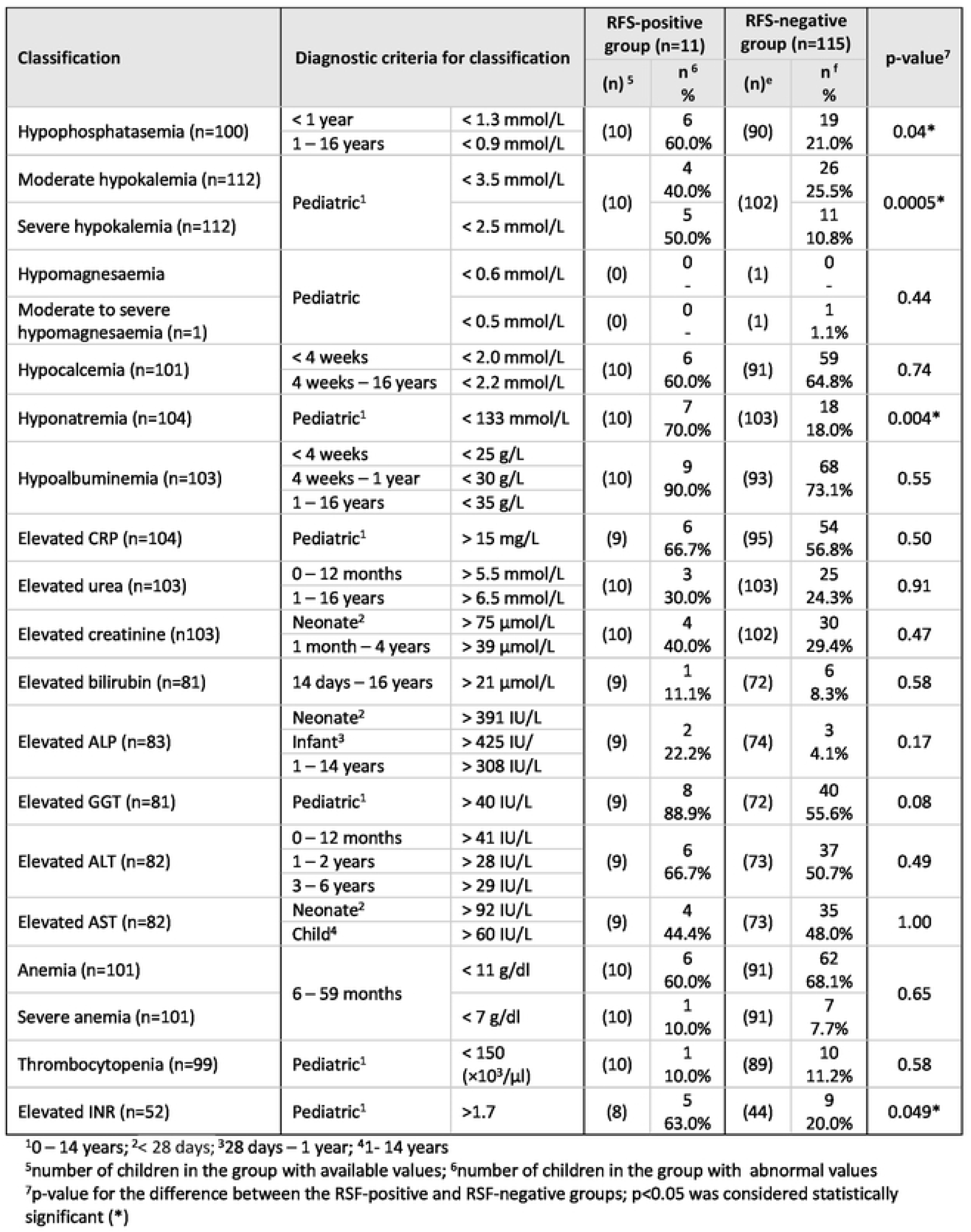
Abnormal biochemical findings on admission (n=126)

Table 4 summarizes the clinical signs and symptoms recorded on admission. Complications on admission were very common, and of the 126 children with SAM in this study, 46.8% presented with vomiting, 52.4% with diarrhea, 53.2% with acute gastroenteritis (AGE), 49.2% with edema, 31% with dermatosis, 10.3% with hypoglycemia, 32.5% with hyperglycemia, 1.6% with hypothermia, 36.5% with pneumonia, 42.9% with respiratory complications, 17.5% with sepsis, 5.6% with septic shock, 60.3% with appetite loss, 37.3% with hepatomegaly, 19.8% with oral thrush, 19.8% with HIV infection, 50% with exposure to HIV, 13.5% with tuberculosis and 19.8% with urinary tract infection (UTI), while 22.2% received nasogastric tube feeding. A significantly higher percentage of children who developed RFS presented with diarrhea (p=0.04), dehydration (p=0.02), and UTI (p=0.04) on admission than those that did not. The RFS positive group included significantly fewer children who were HIV exposed (had a negative PCR negative test but whose mother was positive) than the RFS negative group (p=0.03).

**Table 4:** Clinical signs and medical complications on admission.

| Clinical signs and medical complications | Total group (n=126) |  | RFS-positive group (n=11) |  | RFS-negative group (n=115) |  | p-value<sup>2</sup> |
| --- | --- | --- | --- | --- | --- | --- | --- |
|  | n | % | n | % | n | % |  |
| Vomiting | 59 | 46.8% | 7 | 63.6% | 52 | 45.2% | 0.24 |
| Diarrhea | 66 | 52.4% | 9 | 81.8% | 57 | 49.6% | 0.04* |
| AGE | 67 | 53.2% | 8 | 72.7% | 59 | 51.3% | 0.17 |
| Dehydration | 62 | 49.2% | 9 | 81.8% | 53 | 46.1% | 0.02* |
| Edema | 52 | 41.3% | 7 | 63.6% | 45 | 39.1% | 0.20 |
| Dermatosis | 39 | 31.0% | 5 | 45.5% | 34 | 29.6% | 0.31 |
| Hypoglycemia (< 3 mmol/L) | 13 | 10.3% | 3 | 27.3% | 10 | 8.8% | 0.09 |
| Hyperglycemia (>7 mmol/L) | 41 | 32.5% | 4 | 36.4% | 37 | 32.5% | 0.75 |
| Hypothermia | 2 | 1.6% | 1 | 9.1% | 1 | 0.9% | 0.17 |
| Pneumonia | 46 | 36.5% | 4 | 36.4% | 42 | 36.5% | 1.00 |
| Respiratory complications<sup>1</sup> | 54 | 42.9% | 6 | 54.6% | 48 | 41.7% | 0.53 |
| Sepsis | 22 | 17.5% | 4 | 36.4% | 18 | 15.7% | 0.10 |
| Septic shock | 7 | 5.6% | 1 | 9.1% | 6 | 5.2% | 0.48 |
| Loss of appetite | 76 | 60.3% | 8 | 72.7% | 68 | 59.1% | 0.52 |
| Hepatomegaly | 47 | 37.3% | 6 | 54.6% | 41 | 35.7% | 0.33 |
| Oral thrush | 25 | 19.8% | 1 | 9.1% | 24 | 20.9% | 0.69 |
| HIV infection | 23 | 18.3% | 0 | - | 23 | 20.0% | 0.21 |
| HIV exposed | 63 | 50.0% | 2 | 18.2% | 61 | 53.0% | 0.03*<sup>3</sup> |
| Tuberculosis | 17 | 13.5% | 1 | 9.1% | 16 | 13.9 | 1.00 |
| UTI | 25 | 19.8% | 5 | 45.5% | 20 | 17.4 | 0.04* |
| Nasogastric tube feeding | 28 | 22.2% | 4 | 36.4% | 24 | 20.9% | 0.26 |

<sup>1</sup> dyspnea/respiratory distress, respiratory failure, ventilatory dependency
<sup>2</sup>p-value for the difference between the RSF-positive and RSF-negative groups; p<0.05 was considered statistically significant (\*)
<sup>3</sup>inversely associated

## Discussion

Very few studies, and only one other South African study to date, have focused on RFS in children with malnutrition. This retrospective study among children with SAM treated in a South African public hospital setting recorded an incidence of RFS of 8.7% and found that children who developed RFS were significantly more likely to present with hypophosphatemia, hypokalemia, hyponatremia, dehydration, coagulopathy, and UTI on admission and stayed in the hospital for significantly longer than children who did not develop RFS.

In the current study, the incidence of RFS in children admitted to hospital with SAM was lower than the 15% incidence found by Mbethe and Mda [4] in a teaching hospital in Gauteng, South Africa, and the 21% found by Okinyi [7] in Kenya. However, comparing these three studies is complicated because they used different definitions to define the onset of hypophosphatemia. The current study used the definition identified in a recent systematic review as one of the most used, but its accuracy in predicting adverse outcomes has not been tested in children with SAM [4,5]. Therefore, to effectively study associated risk factors and outcomes of RFS in the context of SAM, a universally accepted definition for hypophosphatemia to indicate the onset of RFS, which is currently lacking [6], needs to be developed.

Hypophosphatemia was present in 20% (n=25) of the 126 children in the current study (Table 3), which concurs with other studies that have reported low phosphate levels on hospital admission for children with SAM [9,18]. In agreement with the findings of Mbethe and Mda [4], hypophosphatemia was significantly (p=0.04) associated with RFS in the current study [5]. Notably, the serum phosphate levels were done on admission before feeds had been initiated, whereas RFS usually occurs within five days of starting to refeed. Therefore, this low phosphate might be a prognostic marker for developing RFS. Moreover, several studies have found that low admission phosphate levels are associated with an increased risk of dying [19,20]. Thus, phosphate assessment is indicated in all patients with SAM to allow for proper supplementation and referral [18,20]. However, regular blood tests to detect a dropping serum phosphate may not be feasible in many settings where SAM is rife, so it is critical to identify risk factors for RFS.

In the current study, 30 (24%) children with SAM presented with severe hypokalemia and 25 (20%) with hyponatremia on admission. According to previous studies, hyponatremia and hypokalemia are the common electrolyte disturbances in malnourished children, made worse by diarrhea, vomiting, and dehydration [21,22]. In addition, hypokalemia and hypomagnesemia frequently occur in children with SAM because of muscle loss and kidney dysfunction due to reductive adaptations during starvation [1,18]. Diarrhea [4,19] and dehydration [18] are also commonly associated with RFS or hypophosphatemia. In the current study, about half of the children presented with diarrhea and/or dehydration on admission, while children who developed RFS during their hospital stay presented with a significantly higher presence of diarrhea (p=0.04), dehydration (p=0.02), severe hypokalemia (p<0.001) and/or hyponatremia (p=0.004) on admission than those who did not.

Coagulopathy has not been described as a risk factor for RFS. However, a report by the WHO identified it as a poor prognostic factor in children admitted with kwashiorkor [23]. De Maayer and Saloojee found that having a prolonged clotting time (INR >1.7) conferred the highest risk of death among children with SAM [24]. In the current study, a significantly higher percentage of children in the RFS-positive group presented with an INR >1.7 on admission than the children in the RFS-negative group (p=0.049), which might reflect the severity of SAM, but further research is required.

Other clinical features commonly associated with RFS or hypophosphatemia are edema [4,18,19] and dermatosis [4,18], which also commonly occur in children with SAM [1]. In the current study, 41% of the children presented with edema on admission. However, although the prevalence of edema was higher in the RFS-positive group than in the RFS-negative group (63.6% vs 31.9%), the difference did not reach statistical significance. Similarly, the prevalence of dermatosis, reported in 31% of the children, was not significantly different between the two groups.

Urinary tract infections are common in malnourished children, and the risk of UTI increases with the severity of malnutrition [25]. Most studies of children with SAM have been conducted in inpatient facilities and have reported a high prevalence of UTI; South Africa reporting among the highest at up to 42% [25]. In the current study, UTI was present in around 20% of the children on admission and was significantly more prevalent (p=0.04) in the group that developed RFS than in those that did not.

In the Kenyan study [7], the prevalence of RFS was significantly associated with HIV infection (but not with dehydration status), while HIV infection was found in a South African study to be associated with an increased risk of death in children with SAM and RFS [24]. The current study in which 18.3% of the children were HIV positive and 50% HIV exposed did not confirm the association with RFS. Moreover, for reasons that are not currently clear, RFS developed significantly more in children who were not HIV exposed than those who were.

The mortality rate amongst the hospitalized South African children with SAM was 20% to 30% in 2012 [26]. Mbethe and Mda [4] reported a mortality rate of 9.5% and noted that most of the children in their study who died had developed RFS. Mortality rates in the current study were 15.1%: 18.2% and 14.5% in the children who developed RFS and those that did not, respectively. However, this may not be a true representation of the study population as 91.7% (n=24) of hospital files of children who demised could be obtained, compared to only 37.7% (n=124) of hospital files for those who were discharged from hospital, indicating retrieval bias. The true mortality in both groups might thus be lower. The only other South African study on RFS in children with SAM reported that 6% of children who developed RFS died [4]. RFS also resulted in a significantly longer duration of hospitalization (18 vs 12 days, p=0.003), which is similar to observations in older adults [12]. The average weight gain during hospitalization for the children with RFS was 0.9 kg (or 7.7 g/kg/day) and 0.5 kg (7.2 g/kg/day) for those without RFS. These differences were not statistically significant, and the trend for higher weight gain in the RFS-positive group may have been attributed to the longer length of hospitalization.

## Data Availability

All relevant data are within the manuscript and its Supporting Information files.

## Limitations

A limitation of this study was the difficulty in obtaining files from the hospital archives, producing a small sample size with potential retrieval bias. In addition, the researchers had to rely on the information recorded in the hospital files and records completed by others. Thus, there was no way of knowing if measurement errors such as inaccurate weighing, measuring of length/height, and MUAC had been made by medical personnel on admission or if errors or omittance occurred when the data was originally captured in the hospital files. Nevertheless, the sample size and findings compare well with similar studies in Africa [4,7].

## Conclusion and recommendations

Refeeding syndrome likely contributes to the high mortality rates experienced in children with SAM, although no direct association with mortality was found in the current study. The association between hypophosphatemia and mortality in children with SAM has been demonstrated previously [18,27]. Therefore, children at risk for developing RFS need to be identified early and monitored closely. The authors recommend that children with SAM and hypophosphatemia, hypokalemia, hyponatremia, INR >1.7, diarrhea, or dehydration need a cautious approach to feeding, regular electrolyte monitoring, and replacement of deficient nutrients.

However, it must be noted that there is a dearth of research supporting preventive or treatment strategies for RFS in SAM, which was highlighted by a recent systematic review [18]. Manary et al. reported that their systematic search did not find studies comparing nutritional strategies in the transition phase of feeding in SAM [28]. Therefore, as Tickell and Denno [2] argued, robust evidence from well conducted trials is needed to confirm or refute current, expert-opinion-based treatment guidelines, decrease mortality, and improve patient outcomes.

## References

1. WHO. Updates on the management of severe acute malnutrition in infants and children [Internet]. Geneva: World Health Organization; 2013. Available from: http://www.who.int/nutrition/publications/guidelines/updates_management_SAM_infantandchildren/en/

2. Tickell KD, Denno DM. Inpatient management of children with severe acute malnutrition: A review of WHO guidelines. Bull World Health Organ [Internet]. 2016 [cited 2019 Mar 27];94:642–51. Available from: https://www.who.int/bulletin/volumes/94/9/15-162867.pdf

3. Friedli N, Stanga Z, Sebotka L, Culkin A, Kondrup J, Laviano A, et al. Revisiting the refeeding syndrome: Results of a systematic review. Nutrition [Internet]. Elsevier Science; 2017 [cited 2019 Aug 7];35:151–60. Available from: http://dx.doi.org/10.1016/j.nut.2016.05.016

4. Mbethe AP, Mda S. Incidence of refeeding syndrome and its associated factors in South African children hospitalized with severe acute malnutrition. Iran J Pediatr [Internet]. 2017;27:e8297. Available from: https://dx.doi.org/10.5812/ijp.8297

5. Crook MA. Refeeding syndrome: Problems with definition and management. Nutrition [Internet]. Elsevier Inc.; 2014;30:1448–55. Available from: http://dx.doi.org/10.1016/j.nut.2014.03.026

6. da Silva JS V., Seres DS, Sabino K, Adams SC, Cober MP, Evans DC, et al. ASPEN consensus recommendations for refeeding syndrome. Nutr Clin Pract [Internet]. 2020;35:178–95. Available from: https://doi.org/10.1002/ncp.10474

7. Okinyi LK. The prevalence of refeeding syndrome among children with severe acute malnutrition: An observational study in Kenyatta Natalional Hospital, Kenya. Gastroenterol Hepatol J [Internet]. 2018;1:69–72. Available from: http://rpdemos.net/clients/hendunadmin/admin/uploads/source/GHJ/GHJ-18-2-119.pdf

8. Yoshimatsu S, Hossain MI, Islam MM, Chisti MJ, Okada M, Kamoda T, et al. Hypophosphatemia among severely malnourished children with sepsis in Bangladesh. Pediatr Int. 2013;55:79–84.

9. Hother A-L, Girma T, Abdissa A, Molgaard C, Michaelsen KF, Briend A, et al. Serum phosphate and magnesium in children recovering from severe acute undernutrition in Ethiopia: An observational study. BMC Pediatr [Internet]. BioMed Central; 2016;16:1–9. Available from: http://dx.doi.org/10.1186/s12887-016-0712-9

10. Namusoke H, Hother A, Rytter MJH, Kæstel P, Babirekere-iriso E, Girma T, et al. Changes in plasma phosphate during in-patient treatment of children with severe acute malnutrition: an observational study in Uganda. Am J Clin Nutr [Internet]. 2016;103:551–8. Available from: http://ajcn.nutrition.org/content/103/2/551.full.pdf+html

11. Rytter MJH, Babirekere-Iriso E, Namusoke H, Christensen VB, Michaelsen KF, Ritz C, et al. Risk factors for death in children during inpatient treatment of severe acute malnutrition: A prospective cohort study. Am J Clin Nutr [Internet]. American Society of Clinical Nutrition; 2017;105:494–502. Available from: https://pubmed.ncbi.nlm.nih.gov/28031190/

12. Friedli N, Baumann J, Hummel R, Kloter M, Odermatt J, Fehr R, et al. Refeeding syndrome is associated with increased mortality in malnourished medical inpatients. Medicine (Baltimore) [Internet]. Lippincott Williams and Wilkins; 2020 [cited 2020 Aug 20];99:e18506. Available from: https://journals.lww.com/10.1097/MD.0000000000018506

13. Friedli N, Odermatt J, Reber E, Schuetz P, Stanga Z. Refeeding syndrome: Update and clinical advice for prevention, diagnosis and treatment. Curr Opin Gastroenterol. 2020;36:136–40.

14. Meyer R, Marino L. Nutrition in critically ill children. In: Shaw V, editor. Clin Paediatr Diet. Fifth edit. Oxford, UK: John Wiley & Sons, Ltd; 2020. p. 80–96.

15. O’Connor G, Nicholls D. Eating disorders. In: Shaw V, editor. Clin Paediatr Diet. Fifth. Oxford, UK: John Wiley & Sons, Ltd; 2020. p. 393–404.

16. Johnson T. Eneral Nutrition. In: Shaw V, editor. Clin Paediatr Diet. Fifth. Oxford, UK: John Wiley & Sons, Ltd; 2020. p. 52–63.

17. Marik PE, Bedigian MK. Refeeding hypophosphatemia in critically ill patients in an intensive care unit. Arch Surg [Internet]. 1996;131:1043–7. Available from: https://www.ncbi.nlm.nih.gov/pubmed/8857900

18. Manary MJ, Hart A, Whyte MP. Severe hypophosphatemia in children with kwashiorkor is associated with increased mortality. J Pediatr. 1998;133:789–91.

19. Freiman I, Pettifor JM, Moodley GM. Serum phosphorus in protein energy malnutrition. J Pediatr Gastroenterol Nutr. 1982;1:547–70.

20. Swanson LC. Prognostic factors in children with severe acute malnutrition at a tertiary hospital in Cape Town, South Africa [Internet]. Stellenbosch University Fa; 2014. Available from: http://scholar.sun.ac.za

21. Raza M, Kumar S, Ejaz M, Azim D, Azizullah S, Hussain A. Electrolyte imbalance in children with severe acute malnutrition at a tertiary care hospital in Pakistan: A cross-sectional study. Cureus. 2020.

22. Gangaraj S, Das G, Madhulata S. Electrolytes and blood sugar changes in severely acute malnourished children and its association with diarrhoea and vomiting. Int J Pharm Sci Invent [Internet]. 2013;2:33–6. Available from: http://www.ijpsi.org

23. WHO. Serious childhood problems in countries with limited resources: background book on management of the child with a serious infection or severe malnutrition [Internet]. Geneva; 2004. Available from: https://apps.who.int/iris/bitstream/handle/10665/42923/9241562692.pdf?sequence=1

24. De Maayer T, Saloojee H. Clinical outcomes of severe malnutrition in a high tuberculosis and HIV setting. Arch Dis Child [Internet]. 2011;96:560–4. Available from: http://www.ncbi.nlm.nih.gov/pubmed/21310895

25. Jones KDJ, Berkley JA. Severe acute malnutrition and infection. Paediatr Int Child Health. 2014;34:S1–29.

26. Department of Health. Integrated Management of Children With Acute Malnutrition in South Africa Operational Guidelines 2015 [Internet]. Pretoria,South Africa: National Department of Health Suggested; 2015. Available from: http://www.rmchsa.org

27. Kimutai D, Maleche-Obimbo E, Kamenwa R, Murila F. Hypo-phosphataemia in children under five years with kwashiorkor and marasmic kwashiorkor. East Afr Med J. 2009;86:330–6.

28. Manary M, Trehan I, Weisz A. Systematic review of transition phase feeding of children with severe acute malnutrition as inpatients. WHO. 2012;1–8.

